# Surface Measure of Fertility and Fertility Transition in India, 1970-2022

**DOI:** 10.1101/2025.08.22.25334278

**Authors:** Aalok Ranjan Chaurasia

## Abstract

**Background:** Total Fertility Rate *(TFR)* is universally used measure of fertility transition but has limitations in analysing fertility transition. This paper proposes an alternative measure of fertility and uses the alternative measure to analyse fertility transition in India.

**Methods:** The paper constructs the Surface Measure of Fertility *(SMF)* based on the presentation of age specific fertility rates *(ASFRs)* as an integrated co-axial chart known as radar chart and shows that *TFR* is a special case of *SMF* when fertility is distributed uniformly across the childbearing period. The difference between *TFR* and *SMF* shows how age distribution of fertility inflates *TFR* relative to *SMF*. The trend in *SMF* reflects the transition in fertility stock or fertility quantum.

**Fertility Transition in India, 1970-2022:** Annual estimates of s available from sample registration system reveal substantial decrease in fertility but unevenly across age groups with periods of stagnation, acceleration, and deceleration in the decrease. *TFR* has consistently been higher than *SMF* because of concentration of births at younger ages of the childbearing period. The median age at childbearing fell until around 2013 leading to increase in the age distribution effects on *TFR* relative to *SMF*. After 2011, median age at childbearing increased because fertility decreased in younger ages but increased in older ages. Changes in age distribution of fertility either slowed down or accelerated decrease in *TFR* relative to *SMF*.

**Conclusions:** *TFR* conflates fertility quantum or fertility stock with age distribution effects, leading to inflated estimates. *SMF* provides a one-to-one mapping of s. Comparison of *SMF* with *TFR* reveals age distribution effects on *TFR*. Indian data shows that concentration of births in the younger ages of childbearing period inflated the *TFR* in the country by as much as 15 per cent during the period 2011-2013. *SMF* better reflects true fertility decline and permits decomposition of *TFR* into stock or quantum component and age distribution component. The *SMF* should be integrated into fertility monitoring frameworks along with *TFR* to provide a more nuanced picture of fertility transition.

## Introduction

Fertility transition encompasses a decrease in the level of fertility and change in the age distribution of fertility. Understanding fertility transition, therefore, requires both analysing the trend in the level of fertility and the change in the age distribution of fertility. It is well-known that the probability to conceive varies by the age of the woman. It, generally, increases with age to reach a peak during the early adult years and then decreases with an increasing rate of decrease with the increase in the age (Bendel and Hua, 1978; Larsen and Yan, 2000; Weinstein et al, 1990). In populations in which natural fertility conditions prevail, childbearing starts at the early ages of the childbearing period and continues throughout the childbearing period so that fertility is well-dispersed across the childbearing period (Singh et al, 2015). Fertility starts decreasing when women start regulating their fertility. The first impetus for regulating fertility is the motivation to prevent unwanted births which leads to the reduction in fertility in the older ages of the childbearing period. Birth limitation is argued to be necessary to reduce the level of fertility for controlling population growth. India was the first country in the world to adopt a policy of birth limitation to control population growth, way back in 1952. An implication of birth limitation is the concentration of fertility in the younger ages of the childbearing period which results in a decrease in the mean age of fertility schedule.

The second impetus for fertility regulation emanates from the importance of proper spacing between successive births primarily from the perspective of survival and health of women and children. There is extensive literature to suggest that proper spacing between births contributes significantly to improving the survival probability of children (Rutstein, 2005; Fotso et al, 2012; Molitoris et al, 2019; Bauserman et al, 2020; Kundu et al, 2025). An increase in birth spacing may not directly lead to a reduction in the level of fertility but it affects fertility through what are known as the child survival hypothesis (Taylor et al, 1976) and the child replacement hypothesis (Temkin-Greenere and Swedlund, 1983). An increase in the spacing between successive births leads to an increase in the dispersion in the age distribution of fertility. On the other hand, a decrease in the spacing between successive births leads to the concentration of fertility in selected ages of the childbearing period, especially, in the prime reproductive period (20-29 years).

The third impetus for fertility regulation is the outcome of the intention to delay childbearing which results in postponing producing children to the older ages of the childbearing period through the delay either at the time of entry into the active reproductive period or increasing the spacing between the time of entry into the active reproductive period and the first birth and between successive births or both. The intention to delay childbearing also leads to a shift in the concentration of fertility from younger ages to older ages of the childbearing period (Frejka and Sardon, 2006; Dioikitopoulos and Varvarigos, 2023; Zabak et al, 2023) and induces a change in the age composition of fertility that results in an increase in the mean age of the fertility schedule.

All the three impetuses for fertility regulation leads to marked changes in the age distribution of fertility. It is, therefore, imperative in any analysis of fertility transition that both change in the level of fertility and change in the age distribution of fertility are analysed. However, the analysis of the change in the age distribution of fertility has received only a residual attention in the analysis of fertility transition, especially, in the developing countries presumably because the developing countries, like India, have been preoccupied with reducing fertility to curb the rapid growth of population. There are studies that have analysed concentration of fertility or the change in the age composition of fertility, generally in the developed countries (Vaupel and Goodwine, 1987; Lutz, 1989; Shkolnikov et al, 2007; Giroux et al, 2008). These studies have generally suggested a negative relationship between the level of fertility and the concentration of fertility – a decrease in fertility has been found to be associated with an increase in the concentration of fertility in the younger ages of the childbearing period. Recently, Spoorenberg (2025) has argued that changing age composition of fertility during fertility transition has implications for the developing countries.

The change in the age composition of fertility also matters for the future growth in population, especially when the replacement fertility is achieved. It is well-known that population continues to grow even after the replacement fertility is achieved because of the in-built momentum for growth in the age composition of the population. In India, for example, population is projected to increase to more than 1701 million by the year 2061 according to the medium but the most likely variant of the population projections prepared by the United Nations, despite the achievement of the replacement fertility sometimes during 2019-2021 (United Nations, 2024). Between 1920 and 2051, more than 300 million people is likely to be added to the population of the country because of population momentum. The change in the age schedule of fertility in a manner that leads to an increase in the mean age of fertility schedule can contribute to reducing population momentum and, therefore, has a substantial impact on the future growth of the population (Bongaarts, 1994).

The total fertility rate *(TFR)* is the universally used measure to analyse fertility transition. *TFR* is the sum of the average number of live births borne by women of different ages of the childbearing period (different cohorts) in a calendar year (single period) or the sum of age-specific fertility rates (s) in a calendar year. It is a period measure of the level of fertility. *TFR* has also been conceptualised as the average number of live births borne to women of a specific age (single cohort) during their entire reproductive life (different periods) if they follow a set of *s* that prevail in a given period. In this conceptualisation, *TFR* is the average number of live births borne by a hypothetical cohort of women during their entire reproductive life and, therefore, is termed as the synthetic measure of fertility. It is, however, rare that any real cohort of women ever follows the *s* which prevail in a given period. The two definitions are, however, not equivalent. The first provides the period perspective of the level of fertility while the other provides the perspective of a synthetic cohort.

The *TFR*, however, has problems in measuring the level of fertility either as a period measure or a cohort measure. The use of *TFR* in analysing fertility transition has been criticised on many grounds (Hajnal, 1947; Ni Bhrolchein, 1992; Bongaarts and Feeney, 1998). These criticisms include the problems posed by the impact of changes in the timing of childbearing or the age distribution of fertility on *TFR;* the problems associated with relating the period measure with the cohort measure; the nature and validity of interpreting period measures as measures of ‘hypothetical’ cohorts; and the extent to which fertility measures should embody controls not only for the age but also for other demographic variables. These concerns are related to analysing fertility transition through a cohort perspective and the solutions that have been proposed have been in the form of adjustments in *TFR* to the change in the age distribution of fertility (Brass, 1991; Ryder, 1964; Murphy, 1994; Bongaarts and Feeney, 1998; Kohler and Philipov, 2001; Ortega and Kohler, 2001; Kohler and Ortega, 2002). Using *TFR* to analyse fertility transition through the period perspective is also problematic. **Ni** Bhrolchein (1992) has argued that analysing fertility transition through the period perspective holds the greater promise and has the more solid justification. Analysing fertility transition through the period perspective is more relevant in assessing the impact of population policies and fertility regulation efforts than analysing fertility transition through the cohort perspective. *TFR* has serious limitations in analyse fertility transition through either period perspective or cohort perspective.

It may be emphasised here that there is no single measure to capture the level of fertility in the population at a particular point in time as fertility varies by age. This means that the level of fertility in a population at a particular point in time is characterised by a set of *s* only. In other words, the level of fertility in the population is a multidimensional construct and, therefore, is represented by the fertility vector, the elements of which are *s*. Here, the vector is defined as an ordered tuple of real numbers (Apostol, 1967; Heijungs, 2025). It is, however, possible to map a vector quantity on a single metric or scalar quantity through an appropriate mapping or aggregation function and the resulting single metric or the scalar quantity that reflects the multidimensional construct and is popularly termed as the composite or synthetic measure of the vector quantity. There is, however, always some loss of information in mapping a vector quantity on a scaler quantity which is contingent upon the mapping or aggregation function. There is no mapping or aggregation function that transfers all the information contained in a vector quantity to a scalar quantity. The selection of the mapping or aggregation function, therefore, is very crucial in deciding the quality and reliability of the composite or the synthetic measure of the vector quantity (Zhou et al, 2010; Zhou et al, 2006).

From the above perspective, the *TFR* is a composite or synthetic measure that is linked to the fertility vector defined by the set of *s* through the sum as the mapping or the aggregation function. The sum of *s* is the same as the simple arithmetic mean of *s* multiplied by 35, the length of the childbearing period. The limitations of the simple arithmetic mean and hence the sum as the mapping or aggregation function in the construction of composite measures is, however, well-known and have been widely discussed, particularly, in the context of the construction of the human development index (Desai, 1991, Kovacevic, 2010; Klugman et al, 2011, OECD, 2008). There are two main problems. First is the perfect substitutability or full compensability of the simple arithmetic mean which, in the present context, means that low fertility in some ages of the childbearing period is fully compensated by high fertility in other ages of the childbearing period in the calculation of *TFR*. Because of the full compensability, analysing fertility transition in terms of the change in *TFR* is problematic. The change in *TFR* is the algebraic sum of the change in *s*. There may be a possibility that fertility in different ages or different elements of the fertility vector change in opposite directions so that the algebraic sum of the change in *s* or the change in the elements of the fertility vector is zero. There may also be a possibility that *TFR* decreases despite increase in fertility in some ages of the childbearing period because the sum of the increase in fertility in some ages of the childbearing period is less than the sum of the decrease in fertility in other ages of the childbearing period. By the same argument, *TFR* may also increase despite the decrease in fertility in some ages of the childbearing period. The decrease in *TFR* does not necessarily mean that fertility has decreased in all ages of the childbearing period. Similarly, increase in *TFR* does not necessarily mean that fertility has increased in all ages of the childbearing period. The change in *TFR* tells little about how fertility has changed in different ages of the childbearing period or how the age distribution of fertility has changed.

The second problem with the simple arithmetic mean or the sum as the mapping or aggregation function is that it ignores the variation in *s* or the distribution of the elements of the fertility vector so that the mapping of the fertility vector on *TFR* or on simple arithmetic mean of sis not unique. Different fertility vectors having different distribution of its elements have the same *TFR* if the sum, or the simple arithmetic mean of the elements of the fertility vector or *s* remains the unchanged. Fertility transition, however, is not just the change in the level of fertility or the fertility stock. Fertility transition is also related to the change in the age distribution of fertility as it has implications to both deciding the level of fertility and in terms of population momentum. The change in *TFR* or the simple arithmetic mean of *s* does not reflect the change in the age distribution of fertility. From the perspective of fertility transition, it is imperative that the change in any composite measure of the fertility vector must reflect both change in the fertility stock and the change in the composition of the fertility stock or the age distribution of fertility.

Because of the limitations of *TFR* or the simple arithmetic mean of *s* as a composite measure of the level of fertility in the population, the geometric mean of *s* has also been used as the mapping or aggregation function to map the fertility vector on a single metric or the scalar quantity (Chaurasia, 2025; Krzyzanowska et al, 2015). The geometric mean addresses, at least partially, the problem of full compensability associated with the simple arithmetic mean or the sum. The geometric mean also takes into account the age distribution of fertility, albeit implicitly. However, geometric mean also has problems as mapping or aggregating function to map the fertility vector on a single metric (Anand, 2018). The main limitation of the geometric mean is that if fertility in any age of the childbearing period is zero, then the geometric mean of the elements of the fertility vector or *ASFRs* is always zero irrespective of fertility in other ages of the childbearing period. Another problem with the geometric mean is that, although it takes into account the distribution of fertility by age it cannot distinguish between the stock component and the age composition component of the fertility vector. The change in the geometric mean cannot be decomposed into the change in the fertility stock and the change in the age distribution of fertility. The use of geometric mean as the mapping or aggregation function to map a fertility vector on a single metric is, therefore, limited in the analysis of fertility transition. Both simple arithmetic mean, and geometric mean are special cases of the generalised or power mean (Bullen, 2003). When the power of the generalised mean, *α* = 1, it reduces to simple arithmetic mean. When power of the generalised mean, *α* =0 it reduces to the geometric mean. The geometric mean is also the simple arithmetic mean of the logarithm of *ASFRs* and, therefore, overcomes, only partially, the drawbacks associated with the simple arithmetic mean (Mariani and Ciommi, 2022).

The fertility vector can also be mapped on a single metric by using mapping or aggregation functions other than simple arithmetic mean or geometric mean. Rules that guide the selection of the mapping or aggregation function for the construction of composite indicators are discussed elsewhere (OECD, 2008). An important criterion for the selection of the mapping or aggregation function is whether the mapping function is compensatory or non-compensatory. Compensability is a disadvantage in the analysis of fertility transition as fertility in different ages may change in opposite directions. The mapping or the aggregation function should also be such that it establishes one-to-one relationship between the fertility vector and the composite measure so that every fertility vector has a unique composite measure. It is also important that the composite fertility measure should be decomposable so that the change in the composite fertility measure can be attributed to the change in the components which constitute the composite measure - the level and the age distribution of fertility. Such a decomposition has policy and programme relevance. If the change in the age distribution of fertility accounts, for example, 70 per cent of the change in the composite fertility measure, then there is little relevance of a policy that is directed towards reducing the level of fertility. In such a scenario, the policy should be directed towards modifying the age distribution of fertility in a manner that is conducive to accelerating fertility transition. It is well known that fertility transition is associated with the transition in both fertility stock and age distribution of fertility because of the transition in the motivation for fertility regulation from limiting births to first proper spacing between successive births and then to postponing childbearing to the older ages of the childbearing period.

This paper has two objectives. The first objective of the paper is to construct a composite measure of fertility that is based on the fertility surface approach. The fertility surface approach of mapping the fertility vector on a single metric is different from the conventional linear and multiplicative approaches of mapping a vector quantity on a scalar quantity. This approach accounts for both fertility stock and age composition of fertility to uniquely characterise the fertility vector or a set of *s* by a single metric. The composite measure of fertility constructed in this paper equals the *TFR* only when fertility is uniformly distributed across the childbearing period or *s* are the same for all ages of the childbearing period. Otherwise, the proposed composite measure of fertility is always less than *TFR* and the difference between *TFR* and the proposed composite fertility measure reflects how *TFR* inflates the level of fertility in the population.

The second objective of the paper is to analyse fertility transition in India during the period 1970-2022 using the proposed composite fertility measure. India has now achieved the replacement fertility *(TFR*=2.1), around 10 years later than the target date set in the National Population Policy 2000 (Government of India, 2000). The analysis shows how the deviation in the age distribution of fertility from the uniform distribution has inflated the *TFR* at the given level of fertility as measured through the proposed composite measure of fertility. The findings of the analysis are important from the policy perspective as the push to limit births at the cost of proper spacing between successive births in the official fertility control efforts in India appears to have resulted in the concentration of fertility in the younger ages of the childbearing period.

The paper is organised as follows: The next section of the paper describes the conceptualisation of the fertility surface and outlines the construction of the surface measure of fertility *(5PM)* as the unique representation of the fertility surface along with its basic properties. The third section of the paper analyses fertility transition in India and shows how fertility transition based on composite fertility measure differs from fertility transition based on *TFR*. The fourth section analyses fertility transition in selected states of India for which annual estimates of age-specific fertility rates are available for the period 1970-2022. The last section of the paper summarises the main findings of the analysis and advocates the case of analysing fertility transition in terms of the composite fertility measure instead of *TFR* so as to account for both transition in both level and age composition of fertility.

### Surface Measure of Fertility

For the sake of simplicity, let the childbearing period (15-49 years) is divided into conventional seven quinquennials age groups 15-19,…,45-49 years and are indexed by i = 1, …, 7 respectively. The same approach can be adopted when single year fertility rates are available. Let, there are *w*_*i*_ women in the age group *i* and *n*_*i*_ ≤ *w*_*i*_ of them deliver a live birth in a calendar year or in a given period. Then the probability of delivering a live birth in a calendar year or in a given period by women of the age group *i, p*_*i*_, is given by

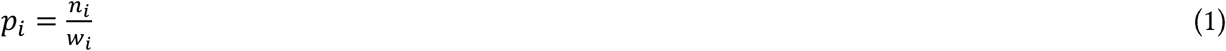

*p*_*i*_ ranges from 0 to 1. If no woman of the age group *i* delivers a live birth in the calendar year, *p*_*i*_=0. If all women of age group *i* deliver a live birth in the calendar year, *p*_*i*_ = 1. The probability vector 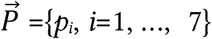, then characterises the fertility profile of the population in terms of the probability of delivering a live birth in a calendar year by women of different age groups.

The vector 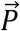 can be visualised as a 7-axes co-axial chart, with each axis of the chart representing the probability of delivering a live birth in a calendar year by women of different age groups as shown in the figure 1. Thus, OA in the figure represents *p*_*1*_, OB represents *p*_*2*_, and so on so that OG represents *p*_*7*_. By connecting points A with B, B with C, C with D, D with E, E with F, F with G, and G with A, the polygon ABCDEFG is constituted which represents the vector of probabilities 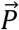 and which uniquely characterises the probability of delivering a live birth in a calendar year by women of different age groups of the childbearing period. The polygon ABCDEFG can be characterised in terms of its size and shape. The size of the polygon may be measured in terms of its area while its shape can be characterised in terms of the deviation of the polygon from regularity. When the polygon is regular, its area is the maximum.

**Figure 1:**
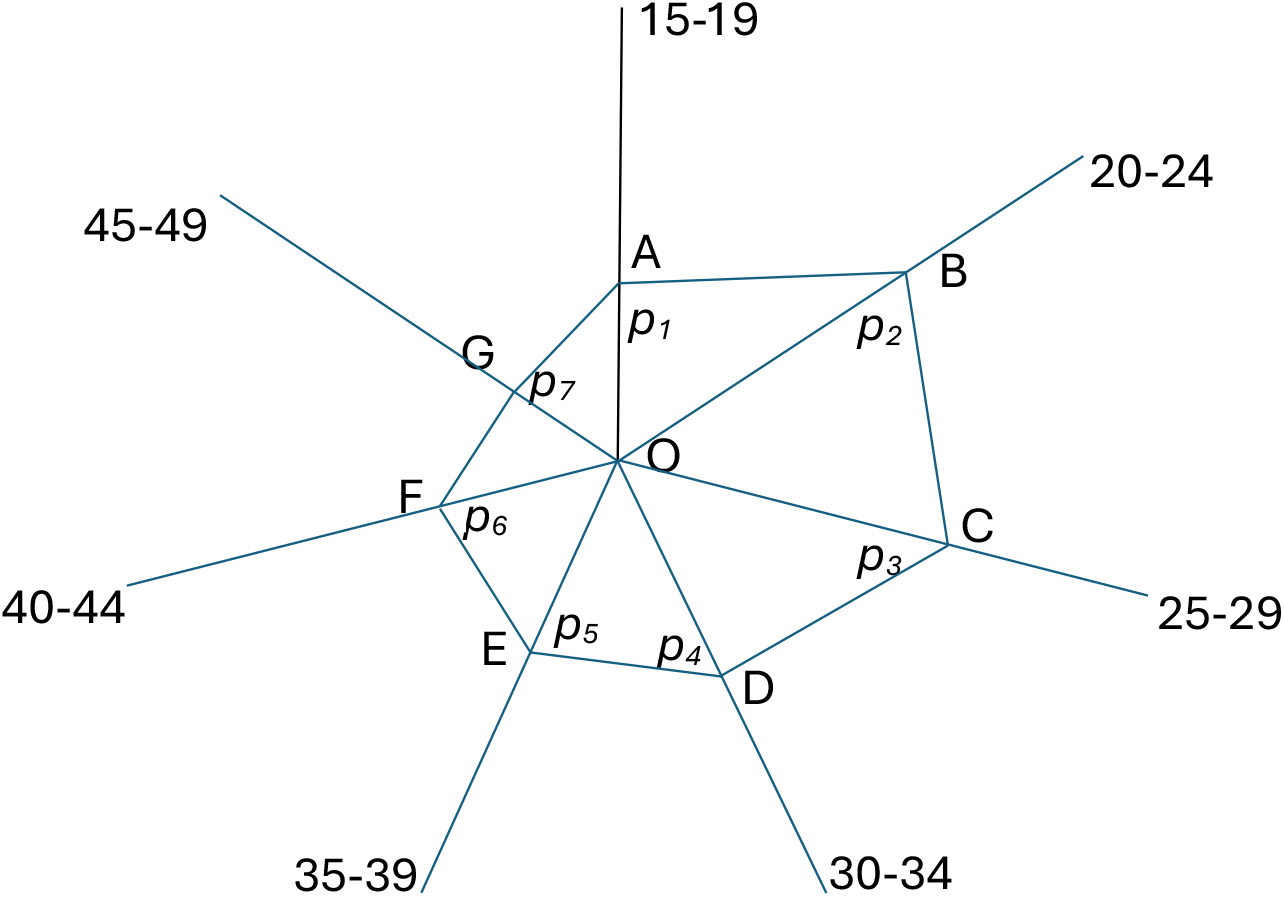
Integrated, co-axial visualisation of the vector of probabilities 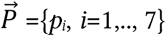. Source: Author

The polygon ABCDEFG comprises of seven triangles OAB, OBC, OCD, ODE, OEF, OFG, and OGA so that the area of the polygon ABCDEFG is the sum of the area of the seven triangles. The area *A* of the polygon ABCDEFG of the figure 1, therefore can be calculated as

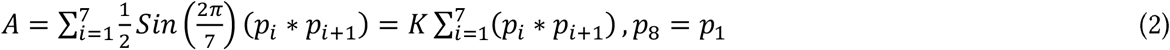

where

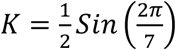

is a constant. When *p*_*i*_= 1 for all *i* =*l*,.., 7, the area of the polygon ABCDEFG is given by

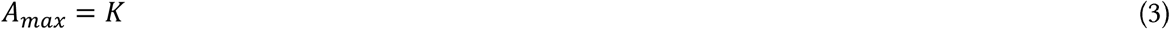

which means that the quantity

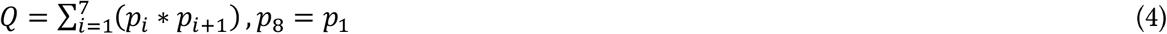

measures the area of the polygon ABCDEFG which ranges from 0 to 1. It can, therefore, be used as a single metric to reflect the area of the polygon or to serve as a single metric to represent the probabilities vector 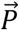.It is also obvious that there is one-to-one mapping or correspondence between the vector quantity 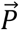 and the scalar quantity *Q*. The scalar quantity *Q* may be taken as the composite measure of population fertility characterised by the probability vector 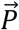.

Three concerns have been raised in using the area of the polygon or the filled up area of the figure 1 as a composite measure of any multidimensional construct or a vector quantity. The first is related to the order of the introduction of different axes in the co-axial chart, It has been shown that the shape and the area of the filled up portion of the co-axial chart varies by the order of introduction of different axes when the number of elements in the vector is more than three (Feldman, 2013; Albo et al., 2016; Heijungs, 2022). The second concern is that the filled up area of the chart is the sum of the area of the triangles that constitute the polygon. The area of the triangle is proportional to the square of the sides of the triangle not to the sides of the triangle. This means that a change in the sides of the triangle results in a larger change in the area of the triangle and hence in the filled-up area of the chart (Zhou, nd). In the present context, this implies that the change in the quantity *Q* exaggerates the change in the probability vector 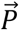. The third concern, on the other hand, is that if the unit of measurement of different elements of the vector is different, then it is necessary to normalise the elements otherwise interpretation of the polygon in terms of its shape and size is meaningless.

In the above context, it may be pointed out that the vector 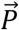 of the probabilities *pi*, is an ordered tuple of non-negative numbers in which the position of each element *(p*_*i*_) is fixed and is determined by its index value which is the age of the woman. This means that the order of introducing different axes in the co-axial chart is always fixed - it begins with *i*=1 and ends with *i*=7 where *i* is the index number. Therefore, there is no arbitrariness in the introduction of the axes in the construction of the polygon. In the present case, the axes in the construction of the co-axial chart are introduced strictly in the order of the index value of the elements of the vector 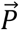 or in order of the age of the woman. The quantity *Q*, therefore, is always unique to the vector 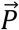 and changes only when the vector 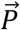 is changed. Two probability vectors can have the same *Q* only when both the elements and the order of the elements are the same. If any of the two are different, *Q* is different.

The second concern related to using the filled up area of the co-axial chart as a composite measure of the probability vector 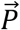 may be resolved by replacing *p;* in equation (4) by an appropriate transformation of *p*_*i*_ so that the change in the area of the triangle is proportional to the change in the sides of the triangle and not to the square of the sides of the triangle. This can be done by replaced *p;* by its Box-Cox transformation with *λ* =l/2 (Box and Cox, 1964). This transformation is the same as the positive square root of *p*_*i*_. The positive square root of any number is a monotonous and continuous transformation of that number. It is similar to the logarithmic transformation of the number, but, unlike the logarithmic transformation, it allows the number to be equal to 0 (Vanella and Deschermeier, 2018). This is particularly relevant here as the probability that a woman delivers a live birth in a calendar year or in a given period may be equal to zero.

Finally, the third concern does not hold in the present case as the elements of the probability vector 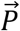 are probabilities and, therefore, they range from the minimum value of zero to the maximum value of 1. There is, therefore, no need of normalising the elements of the probability vector 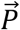. Moreover, the unit of measurement is the same for alls. Now, replacing *p*_*i*_ by its positive square root in equation (4), a composite measure representing the vector 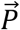 of age-specific probabilities of delivering a birth in a calendar year or in a given period may be defined as

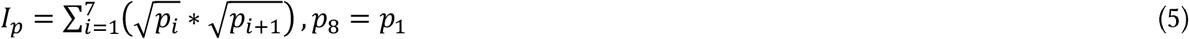

It may be noted that the number of women delivering a live birth in a year is approximately the same as the number of live births delivered in the year. The number of women delivering a live birth in a year is, in fact, marginally lower than the number of live births delivered in the year as some women may deliver more than one live birth in a year. This means that *p*_*i*_, may be very closely approximated by the fertility rate, *f*_*i*_, in the age group *i*. Replacing *p*_*i*_ by *f*_*i*_ and the probability vector 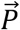 by the fertility vector 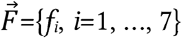, we have

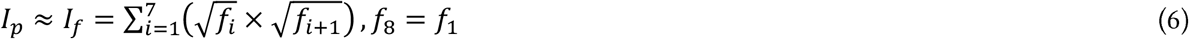

Using equation (6), we can now define the surface measure of fertility *(SMF)*, specific to the fertility vector 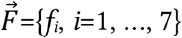 as

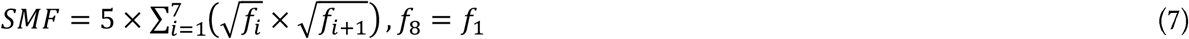

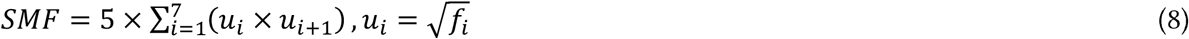

The above conceptualisation visualises fertility in a population at a particular time as seven dimensional construct characterised by the fertility vector *F* which is presented in one integrated co axial chart known by different names including radar chart (Albach and Moerke 1995; Bogan and English 1994; Domptin 1997; Albo et al., 2016; Albuoy et al., 2010; Tague, 2005; Kolence, 1973). This way of presenting multi-dimensional data in one integrated chart was first used by Mayr in 1877 (Mayr, 1877) and is now widely used to visualise progress or performance of any multidimensional construct (Chaurasia and Gulati, 2008; Mosley and Mayer, 1999; Morales-Silva et al., 2020; Porter and Niksair, 2018; Leary et al., 2002; Schutz et al., 1998; Javornik, 2012; Mason et al., 2024; Plantenga and Hansen, 1999; Nigam et al., 2024; Rui et al., 2022; Li and Zhao, 2024; Elbeltagi et al., 2023; Peng et al., 2019; Low et al., 2014; Galagedera and Tan, 2024). The filled-up area of the radar chart serves as a dimensionless, abstract measure of overall progress or performance of the multidimensional construct (Schutz et al, 1998). This approach has also been used to construct the surface measure of human development as an alternative to the conventional HDI (Chaurasia, 2024), a composite index to measure progress towards universal social protection (Chaurasia, 2022), and a composite index to measure nutritional status of children (Chaurasia, 2025). The area of the fertility polygon serves as a measure of the fertility stock in the population - the larger the area the larger the fertility stock. On the other hand, deviation of the shape of the polygon from regularity serves as a measure of the age distribution of fertility.

Notice that equation (7) may be written as

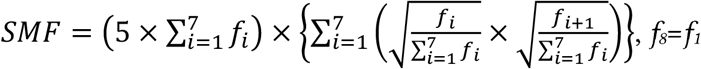

or

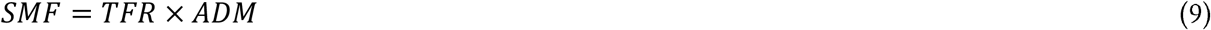

where

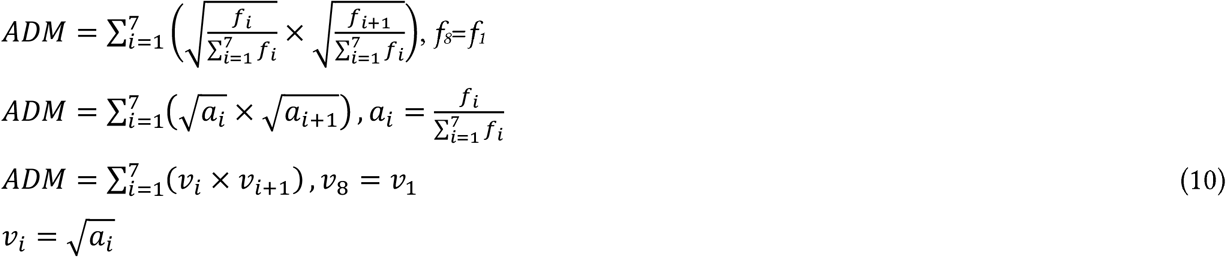

is a measure of the dispersion in the age distribution of fertility - the higher the *ADM* the larger the dispersion in the age distribution of fertility and vice versa. When *f*_*i*_ = *f* for all *i*=l, …,7, *ADM*=1. In this case, *SMF*=*TFR*. Otherwise, *SMF* is always less than *TFR*.

Notice that equation (8) may also be written as

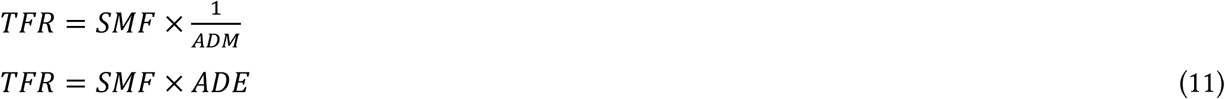

where

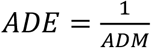

measures the concentration in the age distribution of fertility. When fertility is distributed equally across the childbearing period, *ADE*=1, otherwise *ADE*>1. Equation (11) is of the form

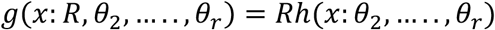

which is the fertility model proposed by Hoems et al (1981). Here *R*=*SMF* represents the fertility stock or the quantum of fertility while *h*(*x*: *θ*_2_, …..,, *θ*_*r*_) = *ADE* is the measure of the deviation of the age distribution of fertility from uniformity. Equation (11) suggests that the higher the *ADE* the higher the deviation of the age distribution of fertility from uniformity and the higher the *TFR* relative to *SME* Equation (11) separates the quantum component (measured by *SMF*) of *TFR* from the age distribution of fertility component (measured by *ADE*) and, therefore, can be used to analyse how change in the age distribution of fertility or timing of births influence the change in *TFR* The area of the fertility polygon reflects the quantum of fertility - the larger the area the higher the fertility quantum or fertility stock and vice versa. On the other hand, the larger the *ADE*, the larger the deviation of the age distribution of fertility from uniformity and, therefore, the larger the effect of the age distribution of fertility on *TFR* Since *ADEM* ≥ 1, the age distribution of fertility always inflates *TFR* relative to the *SME* When *ADE*=1 or when fertility is distributed uniformly across all ages of the childbearing period, *TFR*=*SMF* and, in this case, there is no effect of the age distribution of fertility on *TFR*. In this extreme case, *TFR* measures the stock or the quantum of fertility in the population. It is, however, only exceptional that fertility is distributed uniformly across different ages of the childbearing period as, biologically, fertility varies by age. This implies that *TFR* always inflates the stock or the quantum of fertility implied by the fertility surface measured in terms of *SME* Equation (11) permits decomposing the change in *TFR* into the change in the fertility stock and the change in the age distribution of fertility or the change in the timing of births. Following Kitagawa (1955), the change in TFR can be decomposed as

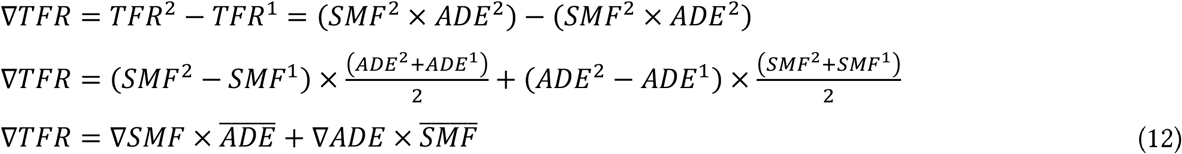

where *TFR*^2^ is *TFR* at time *t*^2^ and *TFR*^1^ is *TFR* at time *t* ^1^ (*t*^2^> *t*^1^). Equation (12) shows that when *ADE* decreases, the decrease in *TFR* is faster than the decrease in *SMF*. However, when *ADE* increases, the decrease in *TFR* is slower than the decrease in *SME* Similarly, when *ADE* decreases, the increase in *TFR* is slower than the increase in *SMF* but when *ADE* increases the increase in *TFR* is faster than the increase in *SMF*. When there is no change in *ADE*, the change in *TFR* is the same as the change in *SMF*. It is, therefore, more appropriate to analyse fertility transition in terms of the trend in *SMF* then in terms of the trend in *TFR*.

The trend in *SMF* can be characterised in terms of the annual proportionate change (*∂*). If *S*^*t*^ denotes *SMF* in the year *t*, then annual proportionate change (*∂*) in SMF, *∂*_*S*_ is calculated as

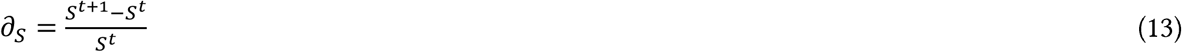

If it is assumed that the trend in *SMF* is linear on a Log scale in the time interval (*t*^*1*^, *t*^*2*^), or if it is assumed that

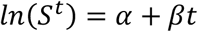

then, it can be shown that

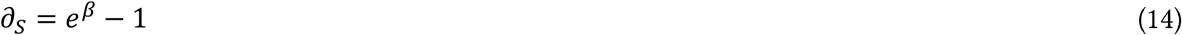

The *∂*_*S*_ indicates the pace of fertility transition. If *∂*_*S*_ = 0 in a time-segment, then there is no change in the pace of fertility transition in the time-segment. When *∂*_*S*_ <0 indicates transition in fertility whereas *∂*_*S*_ >0 signals reversal in transition. When *∂*_*S*_ <0, a decrease in ds indicates acceleration in the transition while an increase in *∂*_*S*_ indicates deceleration in transition. The advantage of measuring the change in terms of *∂* is that it is comparable across scales. It is reasonable to assume that fertility rate in different age groups can change at the same rate, say *x* per cent per year, but it is difficult to assume that fertility in different age groups will change by the same amount on an absolute or arithmetic scale.

The change in SMF is the result of the change in *s* (*f*_*i*_). Let 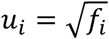, then *∂*_*S*_ can be written as

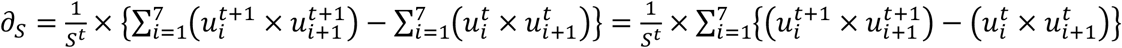

Now

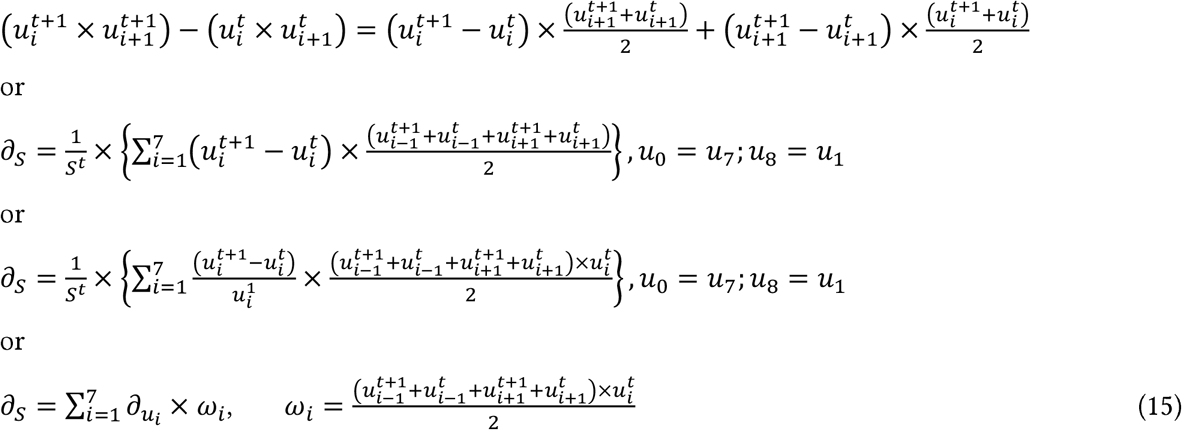

### Fertility Transition in India 1970-2022

We apply the foregoing analytical framework to analyse fertility transition in India during the period 1970-2022. The analysis is based on the estimates of annuals available from the official sample registration system (SRS). The SRS is a large-scale demographic survey that provides annual estimates of key indicators of fertility and mortality at the national and sub-national levels. It was launched on a pilot basis in 1964-1965 and became fully operational in 1969-1970. It is a dual record system which comprises of continuous enumeration of births and deaths in a statistically selected sample units by a resident part time enumerator, and an independent survey every six months by a supervisor. The records of births and deaths obtained from the two independent sources are matched and unmatched and partially matched records are re-verified to obtain an unduplicated count of births and deaths. In the rural areas, the sample unit is either a village or a segment of the village depending upon village population. In the urban areas, the sample unit is a census enumeration block. The sample under the system is replaced every ten years based on the latest population census frame. The current sample is based on the 2011 population census frame as no population census was conducted in 2021. The current sample comprises of 8841 sample units –4,921 rural and 3,881 urban – spread across all states and Union Territories and covers around 8.7 million population (Government of India, 2025). The SRS is the only system in India which provides annual estimates of key fertility and mortality indicators for India and for selected states. Although, registration of births and deaths in India is mandatory under Birth and Death Registration Act of 1969 (Government of India, 1969) which has been amended recently (Government of India, 2023). yet the civil registration system of the country is not designed to estimate fertility and mortality indicators. A major limitation of the civil registration system in India is that it registers births and deaths on *de facto* rather than *de jure* basis. Another problem is incompleteness of birth and death registration. There has been improvement in the registration of births and deaths over the years, but the current completeness is not good enough to estimates fertility and mortality indicators.

Estimates of key indicators of fertility are also available from the National Family Health Survey (NFHS) which was launched by the Government of lndia in 1992-1993 (Government of India, 2022). This survey, however, does not provide annual estimates of indicators of fertility and mortality. The Government of India had also launched the Annual Health Survey Programme in 2010 to generate annually, estimates of fertility and mortality indicators up to the district level (Government of lndia, 2010) but the survey was discontinued in 2013. The SRS is the only source in India that provides annual estimates of indicators of fertility and mortality. The unbroken series of *ASFRs* available through SRS since 1970 provides an opportunity to analyse fertility transition in India over a period of more than 50 years.

Estimates of fertility and mortality indicators, including estimates of *s*, available through SRS are, however, known to be associated with marked annual variations because of errors of unknown origin, although some of these variations may be regarded as real-life phenomena. SRS estimates have also been found to be influenced by the revision of the SRS sample frame. In the past, the SRS sample was replaced after the population census in phases spread over 2-3 years. However, the sample frame was replaced in one go within a year, in 2004 after the 2001 population census, and in 2014 after the 2011 population census (Government of India, 2025).

In view of marked annual variation in the estimates of the indicators of fertility and mortality available through SRS, the standard practice is to use three-years moving average instead of annual estimates available from the system for analysing the trend in fertility and mortality. This approach assumes that the three-years average of the indicator is located at the mid-point of the three-years interval. For example, the average of the interval 1970-1972 is assumed to refer to the year 1971, the mid year of the interval. It has, however, been observed that the three-years average is not the same as the annual estimate available from SRS for the mid-year of the interval. In the present analysis also, we have used the three years moving average of age-specific fertility rates for analysing fertility transition to iron out irregular annual fluctuations in the age-specific fertility rates available through the system.

Figure 2 depicts the decrease in *SMF* and *TFR* in India during the period 1970-2022. The *SMF* or decreased from almost 5.0 to during the period 1970-1972 to around 1.8 during the period 2020-2022 whereas the *TFR* decreased from around 5.2 to slightly more than 2.0 during this period. The *TFR* has always been higher than *SMF* which indicates that the age distribution of fertility has inflated *TFR* relative to *SMF* throughout the period under reference. It may also be seen from the figure that the decrease in both *SMF* and *TFR* has not been consistent and the pace of the decrease in both *SMF* and *TFR* has been different in different time-segments of the period 1970-2022. There are even time-segments in which the decrease in both *SMF* and *TFR* appears to have either stalled or reversed. Another observation of the figure is that the trend in *SFM* has been different from the trend in *TFR*. This is expected as the trend in *TFR* is also influenced by the trend in the timing of births or the change in the age distribution of fertility in addition to the trend in *SMF*. The median age at childbearing decreased from around 28 years during the period 1970-1972 to less than 25.5 years during the period 2011-2013 (Figure 3) but increased rapidly to around 27.8 years by the period 2018-2020. After 2018-2020, the median age at childbearing decreased again to less than 27 years by the period 2020-2022. This means that during the period 1970-2013, fertility in the country got increasingly concentrated in the younger ages of the childbearing period but the trend reversed after 2011-2013. About half of the live births that occurred during the period 1970-1972 in the country were delivered by women younger than 28 years of age whereas about half of the live births that occurred during the period 2011-2013 were delivered by women younger than 25.5 years of age. It is well known that the concentration of fertility in the younger ages of the childbearing period inflates *TFR* relative to the prevailing level of fertility stock or quantum. During 1970-1972, the age distribution of fertility inflated *TFR* relative to *SMF* by less than 5 per cent. This proportion increased to almost 15 per cent during the period 2011-2013 because of the increase in the concentration of fertility in the younger ages of the childbearing period as reflected through the decrease in the median age at childbearing. This proportion, decreased during the period 2011-2020 as the median age at childbearing increased during this period but increased again after 2018-2020. During the period 2020-2022, the age distribution of fertility inflated *TFR* by more than 11 per cent related to *SMF*.

**Figure 2:**
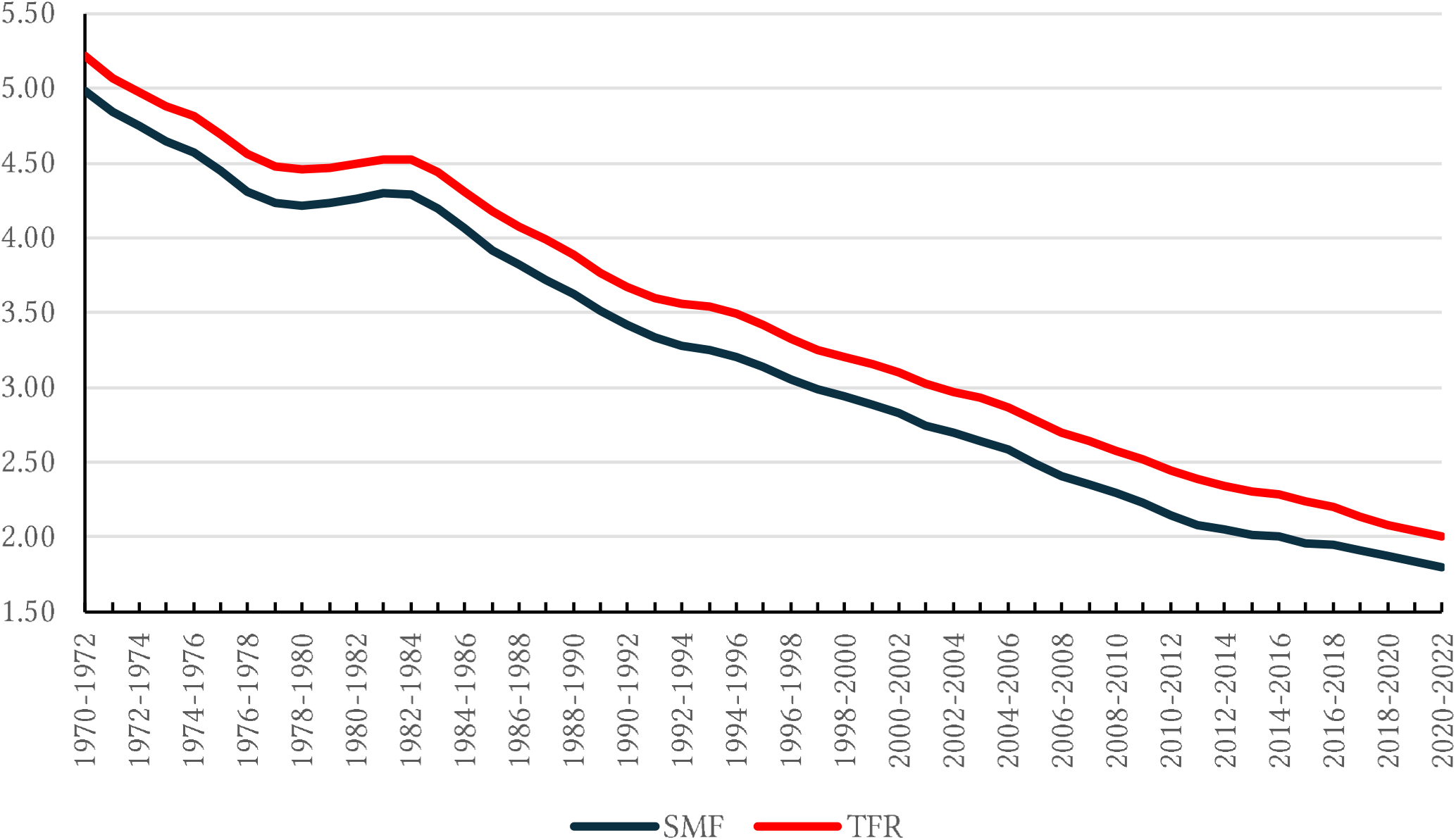
Trend in *TFR*, and *SFM* in India, 1970-2022. Source: Author

**Figure 3:**
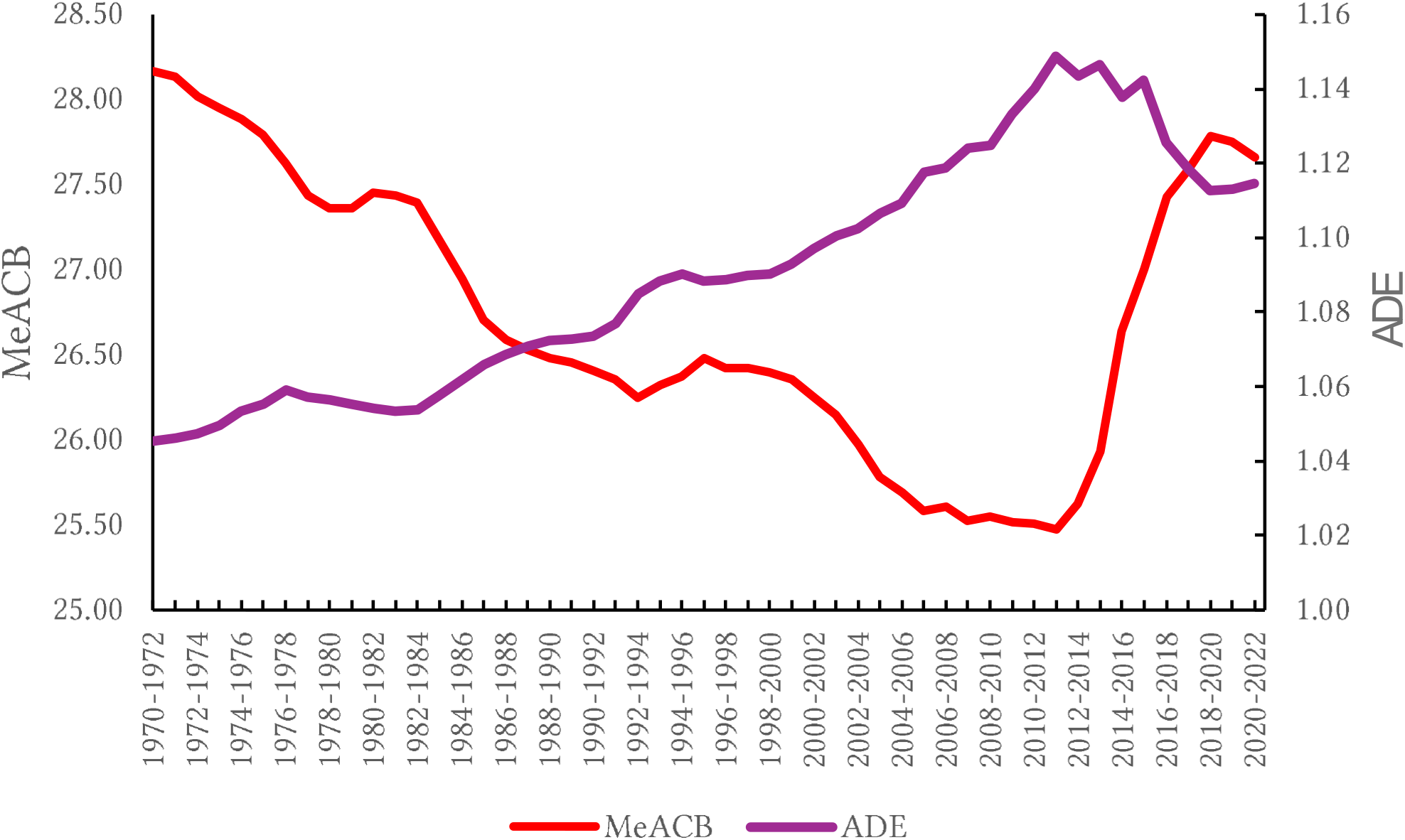
Trend in the mean age at childbearing (*MeACB*) and the effect of the age distribution of fertility on *TFR* (*ADE*) in India, 1970-2022. Source: Author

### Modelling of the Trend in *SMF* and *TFR*

We have used the segmented trend modelling approach to model the trend in *SMF* and *TFR* as the trend in both *SMF* and *TFR* has not been consistent. There are three steps in segmented trend modelling. The first is to test whether there has been a change in the trend. When it is confirmed that there has been a change in the trend, the second step is to identify the time(s) when the trend has changed. If it is found that the trend has changed *k* times, then the entire trend period is divided into *k* +1 time-segments. Finally, the third and the last step is to model the trend in different time-segments separately. If there is no change in the trend, then the segmented trend modelling reduces to the simple trend modelling. The segmented trend modelling is argued to be more appropriate to model the trend in situations where the trend has changed many times (Clegg et al, 2009). The segmented trend modelling best summarises the trend that changes over time (Marrot, 2010).

The most crucial aspect of the segmented trend modelling is to identify the time(s) when there has been a change in the trend so as to divide the trend period into time-segment in which the trend has been consistent. This can be done through visualising the time series. There are different statistical methods that have also been developed for the purpose. The permutation test method (Kim et al, 2000) is regarded as the gold standard. This method uses the sequence of permutation tests to ensure that the approximate probability of the overall Type I error is less than the specified significance level. When there is no change in the trend, “the overall Type I error” is the probability of incorrectly concluding that there has been a change in the trend at least ones during the trend period when, in fact, there is no change in the trend. The permutation method is, however, computationally intensive, especially when the number of times the trend has changed is large.

Alternatively, Bayes factor (Morey et al, 2016) can be used to select the model that best fits the time series. The Bayes factor is the marginalised likelihood of two models, or likelihoods of the two models integrated over the prior probabilities of their parameters (Gill, 2002). The Bayes factor can be thought of as a Bayesian analogue to the likelihood-ratio test, although it uses the marginal likelihood rather than the maximised likelihood. The advantage of the Bayes factor is that it supports the evaluation of the evidence in favour of the null hypothesis, rather than rejecting or not rejecting the null hypothesis (Ly et al, 2020). The computation of the Bayes factor, however, is often challenging depending on the complexity of the model and the hypotheses to be examined (Llorente et al, 2023).

An alternative to estimating the Bayesian factor is based on the Bayesian information criterion (BIC) (Ibrahim et al, 2021). When the data set is large, Bayes factor approaches the BIC as the influence of the priors wanes. The traditional BIC is based on the log-likelihood value which penalises the cost of including extra parameter in the model (Kim et al, 2009). The BIS is computed as

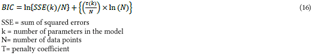

Here the second term on the right hand side is the penalty imposed for including an extra parameter in the model. Using the BIC, that model is selected which has the lowest BIC value.

A number of modifications in the traditional BIC method have been suggested to improve the performance of the method. These include BIC3 method (Kim and Kim, 2016); modified BIC method (Zhang and Siegmund, 2007); and weighted BIC method (Kim et al, 2022). The BIC3 method is a modification of the traditional BIC method. It imposes a harsher penalty for including an extra parameter in the model than the traditional BIC method. The modified BIC method, on the other hand, is based on an asymptotic approximation of the Bayes factor (Zhang and Siegmund, 2007).

The weighted BIC method combines the traditional BIC method and the BIC3 method by using a weighted penalty term that is based on the characteristics of the data. When the duration between the two times when the trend has changed is large, this method assigns a harsher penalty so that the selection rule is based on the BIC3 method. On the other hand, when this duration is small, the method assigns lower penalty so that the selection rule is based on the traditional BIC method (Kim et al, 2022). Use of BIC methods in determining time(s) when the trend has changed is less computationally intensive than the permutation method. Simulation exercises have shown that the methods are nearly equally efficient as the permutation method in modelling the trend that changes over time (National Cancer Institute, 2025). They are, therefore, preferred over the permutation method for segmented trend modelling.

Let the trend has changed *k* times during the trend period so that the entire trend period is divided into *k+*1 time-segments. Then the segmented trend model is defined as:

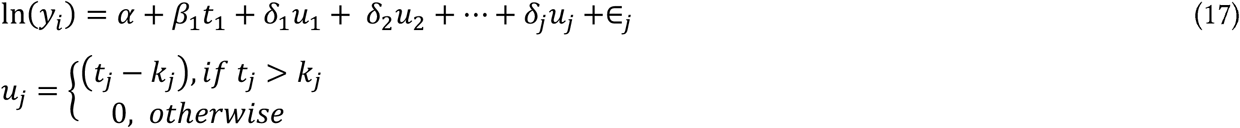

where *y* is the dependent or the study variable and *y;* denotes the value of *y* for the time *t*_*i*_ such that *t*_*1*_*< t*_*2*_*<* … < *t*_*n*_ and *k*_*1*_*< k*_*2*_*<* … < *k*_*j*_ are the times when the trend in *y* has changed.

Actual calculations have been carried out using the Joinpoint Regression Program version 5.0.2 (National Cancer Institute, 2025). The maximum number of times the trend has changed was set to 7 and the weighted BIC (WBIC) method was used to determine the number of times the trend has changed. The grid search method was used to identify the year(s) of the change in the when trend (Lerman, 1980). A grid is created for all possible times of change or their combinations and that time positions are selected which minimises the sum of squared errors. If it is found that the trend has changed *k* times during the trend period, the entire trend period is divided into *k+*1 time-segments, the trend is characterised in each time-segment separately.

Table 1 presents the results of the modelling of the trend in *SMF* and *TFR*. The trend in both *SMF* and *TFR* changed 6 times during the 50 years between 1970-1972 which means that the period 1970-2022 can be divided into 7 time-segments as regards fertility transition. However, the time when the trend in *SMF* changed is not the same as the time-when the trend in *TFR* changed which means that transition depicted by the trend in *SMF* is different from the transition depicted by the trend in *TFR*. The trend in *SMF* reflects the change in the fertility stock or the quantum of fertility whereas the trend in *TFR* reflects the change in both fertility stock and age distribution of fertility.

Table 1 also reveals marked variation in the annual proportionate change (a) in different time segments of the period 1970-2022 in both *SMF* and *TFR*. Fertility transition in India stalled during the period 1977-1984 and again during the period 1881-1996 as the annual proportionate change (a) in both *SMF* and *TFR* was not statistically significantly different from zero during these time-segments. The decrease in both 5MFand *TFR* was the slowest during the period 2010-2022 but was the fastest during the time-segment 2003-2013 in case of *SMF* but during the time-segment 1982-1993 in case of *TFR*. The decrease in *SMF* depicts the change in the fertility stock or fertility quantum as it is not influenced by the change in the age distribution of fertility or the change in the timing of children. If there would have been no change in the age distribution of fertility, the trend in *TFR* would have been the same as the trend in *SMF* and would have depicted the change in fertility stock.

It may also be seen from the table 1 that before the period 2010-2013, both decrease and increase in *TFR* has been slower than the decrease or increase in *SMF*. However, the decrease in *TFR* was faster than the decrease in 5MFafter the period 2010-2022. The decrease in *TFR* is influenced by the change in the age distribution of fertility or the timing of children in addition to the change in *SMF* or the fertility stock. This means that the change in the age distribution of fertility contributed to slow down the change (either decrease or increase) in *TFR* relative to that in *SMF* during the period before 2010-2013 but contributed to accelerate the decrease in *TFR* relative to the decrease in *SMF* during the period after 2010-2013. The median age at childbearing decreased during the period before 2010-2013 which means the increase in the concentration of fertility in the younger ages of the childbearing period. After the period 2010-2013, the median age at childbearing increased which resulted in an increase in the dispersion in the age distribution of fertility.

The average annual proportionate change (Δ) during the trend period may be obtained as the weighted average of the annual proportionate change (a) in different time-segments with weight proportional to the length of the time-segment (Clegg et al, 2009). In other words,

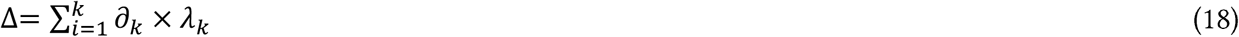

where *λ* _*k*_ is the proportionate length of the time-segment *k* relative to the length of the entire trend period. Δ serves as a summary measure of the trend in different time-segments of the trend period for the purpose of comparison. It is valid even if the model reveals that there are time-segments in which the change is in the opposite direction. Table 1 suggests that, during the entire trend period 1970-2022, *TFR* in India decrease at an average annual rate ofless than 1.9 per cent per year whereas 5MFdecreased at an average annual rate of more than 2 per cent per year which means that the fertility transition in India reflected by the trend in *TFR* has been slower than the fertility transition reflected by the trend in *SMF*. It may, however, be noted that Δ conceals the variation in the annual proportionate change in different time segments. There are time-segments where fertility transition had stalled in the country.

Figure 4 depicts how the fertility surface or polygon has changed in India during the period 1970-2022 in terms of both area and shape reflecting the change in both fertility stock and age composition of fertility. The area of the fertility surface has decreased substantially which reflects the decrease in the average number of live births per woman in a calendar year or the decrease in the fertility stock over the last 50 years. Most of the decrease in fertility surface during the time-segment 2011-2022 has been due to a sharp decrease in fertility in the age group 20-24 years whereas fertility in ages 30 years and above has increased in this time-segment. In absolute terms, the decrease in fertility in the age group 20-24 years in the time-segment 2011-2022 has actually been more than the decrease during the time-segment 1970-2013. The decrease in fertility in this age group has been slower than the decrease in the age group 25-29 years during the time-segment 1970-2005.

**Figure 4:**
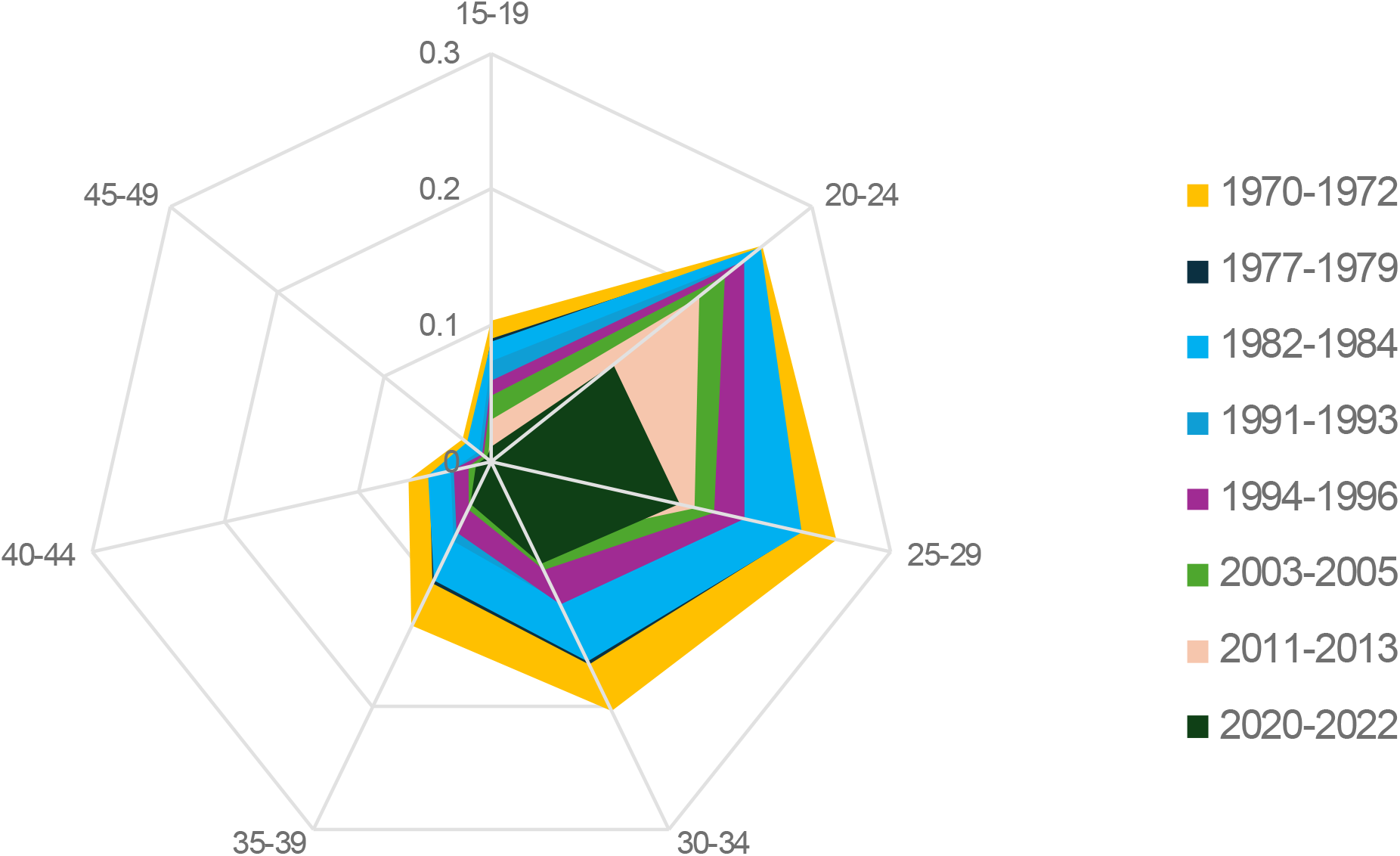
Fertility transition in India, 1970-2022. Source: Author

**Figure 5:**
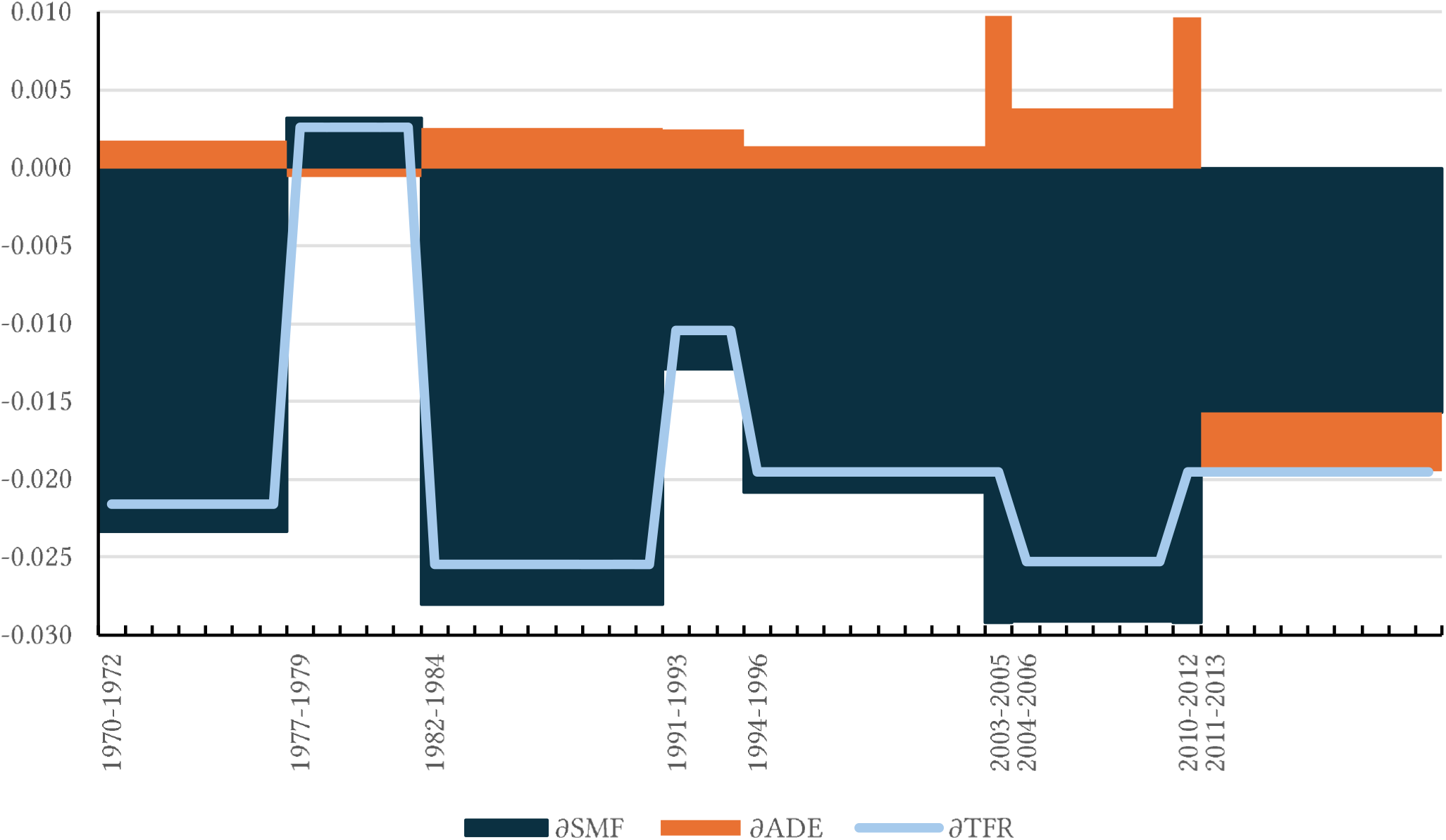
Decomposition of annual proportionate change in *TFR* (*∂* _*TFR*_ xl00) into annual proportionate change in *SMF* (*∂*_*SMF*_ 100) and annual proportionate change in *ADE* (*∂* _*ADE*_ x l00) in India, 1970-2022. Source: Author

The shape of the fertility surface or polygon has also changed over time which indicates the change in the age distribution of fertility or the timing of births. The decrease in the asymmetry in the shape of fertility polygon or the decrease in the skewness of the age distribution of fertility during the period 2011-2022 has been due to both a rapid decrease in fertility in the age-group 20-24 years and a marked increase in fertility in ages 30 years and above. In contrast, the dispersion in the age distribution of fertility appears to have decreased during the period 1970-2013 which means increased concentration in the age distribution of fertility and hence an increase in the skewness of the age distribution of fertility leading to an increase in *TFR* relative to *SMF*.

### Age Distribution Effects of TFR

The age distribution of fertility is not uniform so that it has inflated *TFR* relative to *SMF*. During the period 1970-1972, the measure of uniformity in the age distribution of fertility, *ADM*, was 0.957 which means that the age distribution of fertility inflated *TFR* relative to *SMF* by around 4.5 per cent The *ADM* decreased to 0.870 during the period 2011-2013 which means that the age distribution of fertility inflated *TFR* relative to *SMF* by almost 15 per cent because of the marked decrease in the uniformity in the age distribution of fertility. After 2011-2013, *ADM* increased to almost 0.900 during the period 2018-2020 and has remained virtually unchanged since then so that the age distribution of fertility inflated *TFR* relative to *SMF* by more than 11 per cent during the period 2020-2022. Because of the change in the age distribution of fertility, fertility transition depicted by the trend in *TFR* was slower than fertility transition depicted by the trend in *SMF* during the period 1970-2013. However, fertility transition depicted by the trend in *TFR* during the period 2011-2022 has been faster than fertility transition depicted by the trend in *SMF* during the period 2011-2022 because of the increase in the measure of the age distribution of fertility, *ADM* which resulted in a decrease in the inflation of *TFR* relative to *SMF* because of the age distribution of fertility.

The inflation of *TFR* relative to *SMF* is the result of the deviation of the age distribution of fertility from the uniformity - the larger the deviation the higher the inflation and vice versa. The deviation of the age distribution of fertility from uniformity increases when fertility in high fertility age groups decreases slowly than the decrease in fertility in those age groups in which fertility is low which means concentration of fertility in those age groups in which fertility is high. On the other hand, the deviation of the age distribution of fertility from uniformity decreases when fertility in high fertility age groups decreases more rapidly than the decrease in fertility in age groups in which fertility is low. The deviation from the uniformity also decreases when the decrease in fertility in high fertility age groups is associated with the increase in fertility in age groups having low fertility. When fertility changes in the same proportion in all age groups, there is no change in the deviation of the age distribution of fertility from uniformity.

The segmented trend analysis suggests that the trend in *ADE* or, equivalently, in *ADM* changed three times during the period 1970-2022. During the period 1970-1984, the *ADE* increased at an annual rate of around 0.08 per cent per year. The annual rate of increase in *ADE* increased to 0.21 per cent per year during the period 1982-2005 and to almost 0.43 per cent per year during the period 2003-2013. However, after 2011-2013, the *ADE* decreased at an annual rate of more than 0.43 per cent per year. As a result, the *TFR* inflation effect of the age distribution of fertility increased up to 2011-2013 but decreased thereafter. The increase in inflation effect of the age distribution of fertility contributed to slow down the decrease in *TFR* relative to *SMF* during the period 1970-2013 and this deceleration was quite marked during the period 2003-2013. However, the decrease in inflation effect of the age distribution of fertility on *TFR* during the period 2011-2022 accelerated the decrease in *TFR* relative to *SMF*. If there had been no change in the age distribution of fertility, then the proportionate decrease in *TFR* in different time segments would have been the same as the proportionate decrease in *SMF* and there would have been no change in the inflating effect of the age distribution of fertility on *TFR* relative to *SMF*.

Table 3 decomposes the annual proportionate change (a) in *TFR, ∂_*TFR*_*, in different time-segments of the period 1970-2022 into two components, one attributed to annual proportionate change in *SMF, ∂_*SMF*_*, and other to annual proportionate change in *ADE, ∂_*ADE*_*. If there is no change in the age distribution of fertility in a time-segment, *∂_*ADE*_*=*0* and *∂_*TFR*_*=*∂_*SMF*_* in the time-segment. However, *∂_*ADE*_>0* during the period 1970-1979 and 1982-2013 so that *∂_*TFR*_<∂_*SMF*_* during these time-segments. On the other hand, *∂_*ADE*_<0* during time-segments 1977-1984 and 2012-2022 so that increase in *TFR* during the time-segment 1977-1984 was slower than the increase in *SMF* whereas the decrease in *TFR* during the time-segment 2012-2022 was faster than the decrease in *SMF* during this time-segment. Table 3 shows how the age distribution of fertility has influenced the change in *TFR* in India. The change in the age distribution of fertility as reflected through the change in the shape of the fertility surface either decelerates or accelerates the change in *TFR* relative to the change in *SMF*.

**Table 2:**
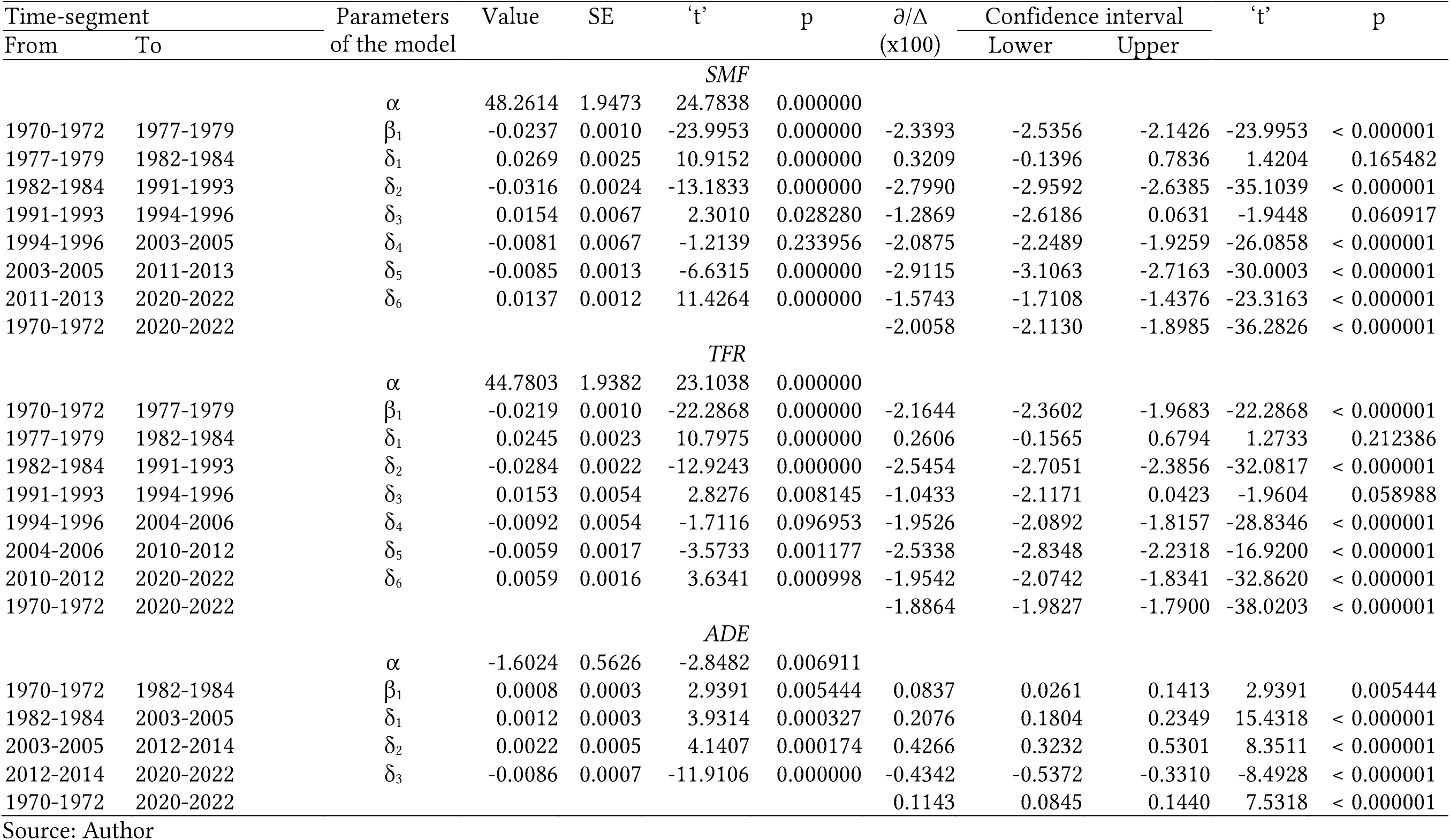
Results of the segmented analysis of the trend in *TFR, SMF* and *ADE* during the period 1970-2022 in India.

**Table 3:** Contribution of *∂*_*SMF*_(x100) and *∂*_*ADR*_(x100) to *∂*_*TFR*_ in different time-segments.

| Time-segment | | $\partial_{TFR}$<br>during the<br>time-segment | $\partial_{TFR}$ attributed to | |
| --- | --- | --- | --- | --- |
| From | To | | $\partial_{SMF}$ | $\partial_{ADE}$ |
| 1970-1972 | 1977-1979 | -0.0216 | -0.0234 | 0.0018 |
| 1977-1979 | 1982-1984 | 0.0026 | 0.0032 | -0.0006 |
| 1982-1984 | 1991-1993 | -0.0255 | -0.0280 | 0.0026 |
| 1991-1993 | 1994-1996 | -0.0104 | -0.0129 | 0.0025 |
| 1994-1996 | 2003-2005 | -0.0195 | -0.0209 | 0.0014 |
| 2003-2005 | 2004-2006 | -0.0195 | -0.0293 | 0.0097 |
| 2004-2006 | 2011-2013 | -0.0253 | -0.0292 | 0.0038 |
| 2011-2013 | 2012-2014 | -0.0195 | -0.0293 | 0.0097 |
| 2012-2014 | 2020-2022 | -0.0195 | -0.0157 | -0.0038 |

Source: Author.

The decomposition results are supported by the change in the median age at childbearing or the age of the women by which half of the live births occur. During the period between 1970-1972 and 1977-1979, the median age at childbearing in the country decreased from around 28.2 years to almost 27.4 years, an average annual decrease of around 0.1 year per year which indicates an increase in the concentration of fertility in the younger ages of the childbearing period. Between 1977-1979 and 1982-1985, there has been virtually little change in the median age at childbearing but, between 1982-1985 and 1991-1993, the median age at childbearing decreased at an average annual rate of almost 0.12 years per year. The median age at childbearing increased marginally between 1991-1993 and 1994-1996 but decreased again between 1991-1996 and 2004-2006 at an average annual rate of 0.07 years per year and between 2004-2006 and 2011-2013 at an average annual rate of 0.03 years per year so that the median age at childbearing of women was the lowest during the period 2011-2013 during the entire reference period. However, after 2011-2013, the median age at childbearing increased very rapidly from around 25.5 years during the period 2011-2013 to around 27.7 years during the period 2020-2022 at an average annual rate of more than 0.24 years per year so that the median age at childbearing increased to almost 27.7 years during the period 2020-2022. A decrease in the median age at childbearing indicates an increase in the concentration of fertility in the younger ages of the childbearing period which results an increase in the inflation of *TFR* relative to *SMF* due to the change in the age distribution of fertility and a slowdown in the decrease in *TFR* relative to the decrease in *SMF*. On the other hand, an increase in the median age at childbearing indicates a decrease in the concentration of fertility in the younger ages of the childbearing period which results in a decrease in the inflation of *TFR* relative to *SMF* due to the change in the age distribution of fertility and contributes to accelerating the decrease in *TFR* relative to *SMF*. The change in the age distribution of fertility influences the decrease in *TFR* but not the decrease in *SMF*. The trend in *SMF* reflects the change in the fertility stock or the fertility quantum whereas the trend in *TFR* reflects the change in both fertility stock and age distribution of fertility.

### Decomposition of the Change in *SMF*

The change in *SMF* is contingent upon the change in s. Figure 6 presents annual proportionate change (*∂*) in fertility rate in different age groups of the childbearing period. The change in s in India has been highly inconsistent across age groups as well as across time. Because of this inconsistency in the change in fertility rate in different age-groups, the contribution of the change in fertility in an age-group to the change in *SMF* has also been highly inconsistent as may be seen from the figure 7 and table 4. There is no age-group in which fertility decreased consistently throughout the 50 years under reference. In all age groups, there are time segments in which fertility increased, instead decreased. Notably, there has, in general, been an increase in fertility in older ages of the childbearing period after the period 2011-2013 but a decrease in the younger ages. As the result, there has been an increase in the dispersion in the age distribution of fertility which has resulted in a decrease in the fertility inflating effect of the age distribution of fertility on *TFR* relative to *SMF*.

**Figure 6:**
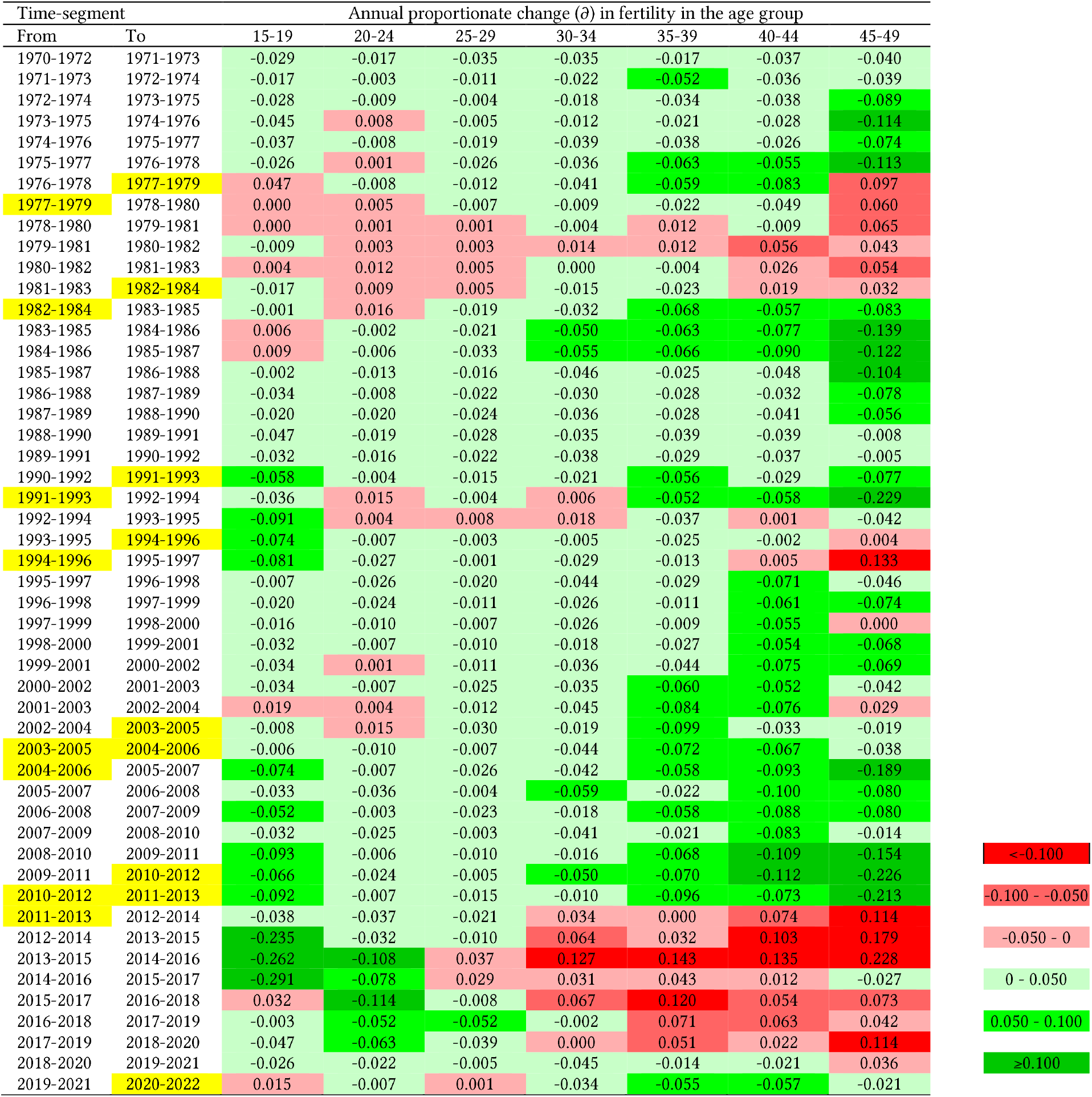
Annual proportionate change *(∂)* ins in India, 1970-2022. Source: Author

**Figure 7:**
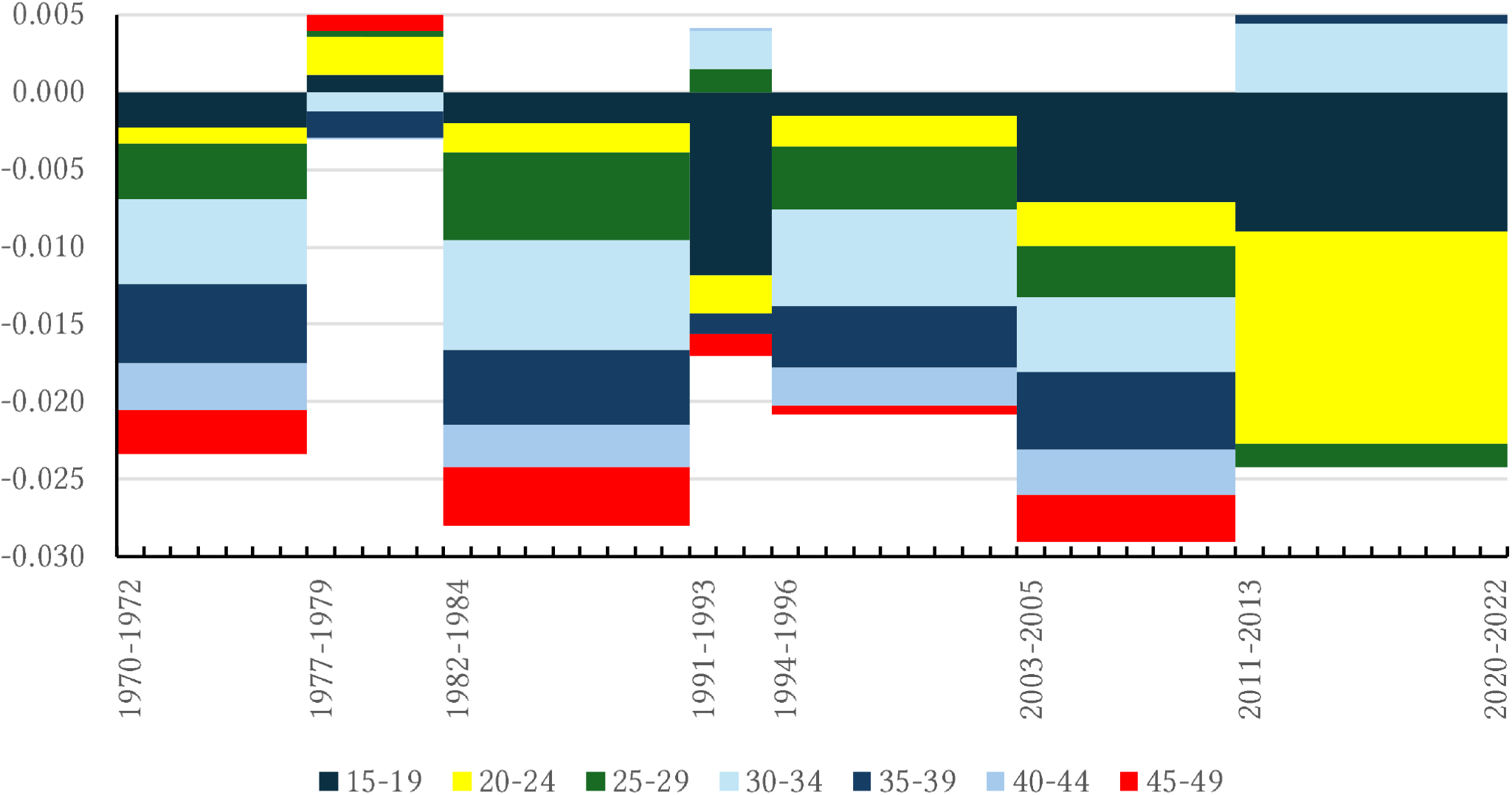
Contribution of the annual proportionate change (*∂*) in s to the annual proportionate change (*∂*) in *SMF* in different time-segments of the period 1970-2022. Source: Author

Table 5 presents the contribution of the annual proportionate change (a) in *ASFRs* to the annual proportionate change (a) in *SMF* in different time-segments of the period 1970-2022 identified through segmented trend analysis. In four time-segments, decrease in fertility in all age groups contributed to the decrease in 5MFwhereas in three time-segments, fertility decreased in some age groups but increased in other age groups so that the decrease in *SMF* slowed down considerably in two time-segments and increased in one time-segment. However, the increase in *SMF* during the period 1977-1984 and the decrease in *SMF* during the period 1991-1996 has not been found to be statistically significantly different from zero which means that the decrease in fertility stagnated in these time-segments.

**Table 5:**
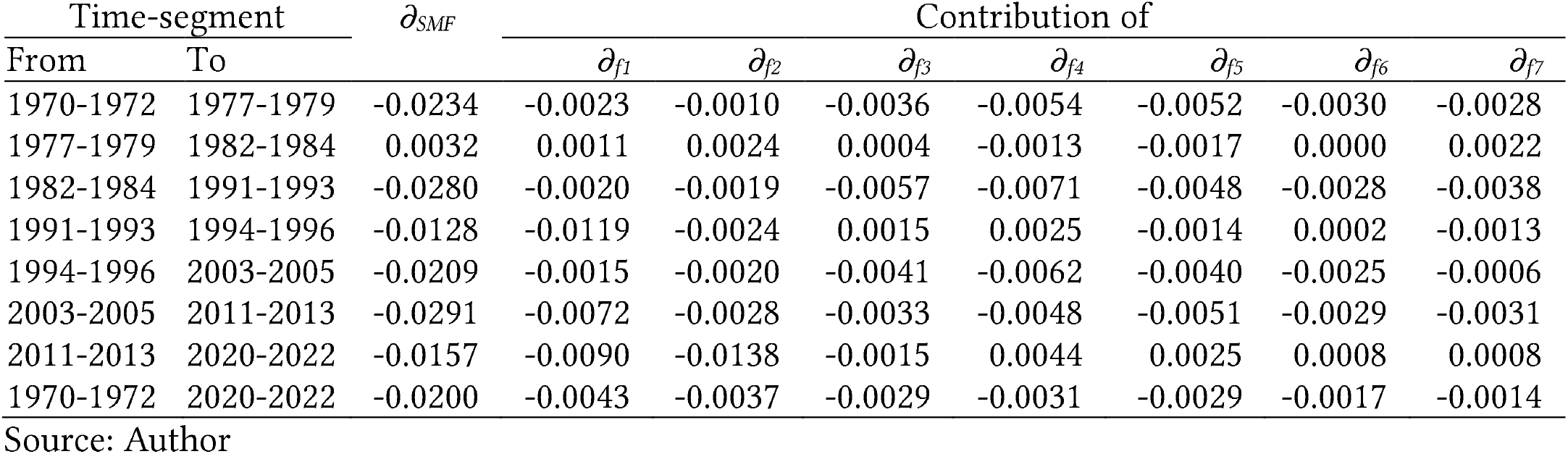
Contribution of annual proportionate change *∂* in fertility in different age-groups to *∂_*SMF*_* in different time-segments.

| Time-segment | | $\partial_{SMF}$ | Contribution of | | | | | | |
| --- | --- | --- | --- | --- | --- | --- | --- | --- | --- |
| From | To | | $\partial_{f1}$ | $\partial_{f2}$ | $\partial_{f3}$ | $\partial_{f4}$ | $\partial_{f5}$ | $\partial_{f6}$ | $\partial_{f7}$ |
| 1970-1972 | 1977-1979 | -0.0234 | -0.0023 | -0.0010 | -0.0036 | -0.0054 | -0.0052 | -0.0030 | -0.0028 |
| 1977-1979 | 1982-1984 | 0.0032 | 0.0011 | 0.0024 | 0.0004 | -0.0013 | -0.0017 | 0.0000 | 0.0022 |
| 1982-1984 | 1991-1993 | -0.0280 | -0.0020 | -0.0019 | -0.0057 | -0.0071 | -0.0048 | -0.0028 | -0.0038 |
| 1991-1993 | 1994-1996 | -0.0128 | -0.0119 | -0.0024 | 0.0015 | 0.0025 | -0.0014 | 0.0002 | -0.0013 |
| 1994-1996 | 2003-2005 | -0.0209 | -0.0015 | -0.0020 | -0.0041 | -0.0062 | -0.0040 | -0.0025 | -0.0006 |
| 2003-2005 | 2011-2013 | -0.0291 | -0.0072 | -0.0028 | -0.0033 | -0.0048 | -0.0051 | -0.0029 | -0.0031 |
| 2011-2013 | 2020-2022 | -0.0157 | -0.0090 | -0.0138 | -0.0015 | 0.0044 | 0.0025 | 0.0008 | 0.0008 |
| 1970-1972 | 2020-2022 | -0.0200 | -0.0043 | -0.0037 | -0.0029 | -0.0031 | -0.0029 | -0.0017 | -0.0014 |

Source: Author

During the period 2011-2022, on the other hand, the decrease in fertility has been confined to the younger ages of the childbearing period only. The decrease in fertility has been very rapid in the age groups 20-24 years and 15-19 years. On the other hand, there has been an increase in fertility in ages 30 years and above during this period with the increase being quite sharp in the age group 30-34 years. As the result, the decrease in *SMF* markedly decelerated during the period 2011-2013 relative to the period 2003-2013. The decrease in *TFR*, however, accelerated during this period because of the increase in the dispersion of the age distribution of fertility.

## Discussion

There is no single metric or indicator to measure fertility in the population at a particular point in time. Fertility varies by the age of the woman and, therefore, fertility in the population can best be characterised by an ordered set of age-specific fertility rates or the fertility vector. The fertility vector can be mapped on a single metric through a mapping operator or aggregation function so that the single metric or the scalar quantity so obtained serves as a composite measure of fertility in the population determined by the fertility vector and fertility transition may be analysed in terms of the change in the composite measure. It is obvious that the value of the composite measure depends upon the mapping operator or the aggregation function used to map the fertility vector on the composite measure. Different mapping operator or aggregation function leads to different composite measures for the same fertility vector. Selection of the mapping operator or the aggregation function, therefore, is critical to constructing a composite measure of that adequately reflects the fertility vector. An important requirement for the selection of the mapping operator or the aggregation function is that the relationship between the fertility vector and the composite measure defined by the mapping operator or the aggregation function must be unique in the sense that two different fertility vectors must not have the same composite measure.

The *TFR*, universally used to measure population fertility and analyse fertility transition, is a composite measure of the fertility vector with the sum or the simple arithmetic mean of the elements of the fertility vector or fertility in different ages of the childbearing period as the mapping operator or the aggregation function. The limitations of constructing a composite measure using the simple arithmetic mean or the sum as the mapping operator or the aggregation function are well-known and well documented. Because of these limitations, the problems associated with analysis of fertility transition in terms of the change in *TFR* are also well documented. The main limitations of *TFR* is that it is sensitive to the age distribution of fertility or the timing of births so that *TFR* comprises of two components - a quantum or stock component and an age composition component. It has been argued that fertility transition should be analysed in terms of the change in fertility stock or quantum of fertility which is not sensitive to the age composition of fertility. There have been attempts to adjust the *TFR* for the effect of the age distribution of fertility to measure the fertility stock or the quantum of fertility but the challenge of analysing fertility transition in terms of the change in *TFR* remains. There has, however, rarely been attempts to construct alternative composite measures of the fertility vector or s that reflect the fertility stock or the quantum of fertility. This is so when there has been extensive research in recent years on the construction of the composite indicators.

This paper has proposed an alternative composite measure of the fertility vector based on the concept of fertility surface which is formed by presenting s as a single integrated, coaxial chart popularly known as the radar chart and the area of the fertility surface serves as a measure of the fertility stock or the quantum of fertility - the larger the area of the fertility surface the higher the fertility stock or the quantum of fertility and vice versa. Based on the concept of the fertility surface, the paper proposes the surface measure of fertility *(SMF)* as a composite measure to reflect the fertility stock or quantum of the fertility vector, and which is unique to the fertility vector. If two fertility vectors are different, then their surface measure of fertility is also different. *TFR* is an especial case of *SMF* when the age distribution of fertility is a uniform distribution or when fertility is the same in all ages of the childbearing period. In this case, the fertility surface is a regular polygon and the deviation of a fertility polygon from the regularity is a measure of the lack of equality or inequality in the age distribution of fertility. The inequality in the age distribution of fertility reflects the concentration of fertility in some ages of the childbearing period which inflates *TFR* relative to *SMF*. Using *SMF* as a measure of fertility stock or fertility quantum, the age distribution effect of *TFR* can be estimated as the proportion by which *TFR* is higher than *SMF*. The change in *TFR* depends upon both change in fertility stock and change in the effect of the age distribution of fertility on *TFR*. Since the area of the fertility surface is independent of the age distribution of fertility, the trend in *SMF* depicts the transition in the fertility stock or the quantum of fertility independent of the age distribution of fertility.

Estimates of *TFR* and *SMF* in India for the period 1970-1972 through 2020-2022 reveals that *TFR* has always been higher than 5MFwhich means that the age distribution of fertility in India always inflated *TFR* relative to *SMF*. This finding is supported by the observation that the median age at childbearing in India has never been more than 28 years which means that fertility in the country has largely been concentrated in the younger ages of the childbearing period. The concentration of fertility in the younger ages of the childbearing period increased during the period between 1970-1972 and 2011-2013 as reflected through the decrease in the median age at childbearing so that the age distribution of fertility inflated *TFR* relative to *SMF* by almost 15 per cent during the period 2011-2013. However, the concentration of fertility in the younger ages of the childbearing period decreased after 2011-2013 because of the decrease in fertility in ages 15-29 years but the increase in fertility in ages 30-45 years leading to a decrease in the proportion by which the age distribution of fertility inflated *TFR* relative to *SMF* to around 11 per cent by the period 2020-2022. The increase in inflation in *TFR* due to increased concentration of fertility in the younger ages of the childbearing period resulted in a deceleration in the decrease in *TFR* relative to the decrease in *SMF* during the period 1970-1972 through 2011-2013 but the decrease in inflation in *TFR* after 2011-2013 led to an acceleration in the decrease in *TFR* relative to the decrease in *SMF*. The fertility transition reflected by the trend in *TFR* has, therefore, been different from the fertility transition reflected by the trend in 5MFbecause of the change in the age distribution of fertility before and after 2011-2013.

An interesting finding of the present analysis is that the age distribution effects on *TFR* in India has been different has been different during the period before 2011-2013 and after 2011-2013. During the period before 2011-2013, the change in the age distribution of fertility contributed to inflate *TFR* because of the increase in the concentration of fertility in the younger ages of the childbearing period. However, after the period 2011-2013, fertility in the younger ages of the childbearing period decreased rapidly but fertility in the older ages increased leading to an increase in the dispersion of fertility and, hence, a decrease in the inflating effect of the age distribution of fertility on *TFR*. One possible reason for the marked decrease in fertility in the younger ages of the childbearing period is the increase in the age at marriage as virtually all fertility in India is confined within the institution of marriage. Data available from SRS suggest that around 12 per cent of women aged 15-19 years in India were married during the period 2011-2013. This proportion decreased to around 3 per cent during the period 2020-2022. Similarly, almost 60 per cent of women aged 20-24 years were married during the period 2011-2013 which decreased to around 34 per cent during the period 2020-2020. The marked decrease in the proportion of married women aged 15-24 years during the period 2011-2022 appears to be the main contributor of the decrease in fertility in these age groups as there has been little change in the fertility of married women during this period. On the other hand, fertility of women aged 30 years and above increased during this period which reflects dilution of fertility regulation efforts in the country and need further investigation.

## Conclusions

There is no single indicator to measure population fertility as fertility varies by age. Composite indicators are, therefore used to measure population fertility and to analyse fertility transition. The construction of the composite measure, therefore, matters. *TFR* is a composite measure of fertility which is universally used to measure fertility and analyse fertility transition. The limitations of *TFR*, however, are well-known. Reliance on *TFR* alone in analysing fertility transition may, therefore, be misleading because the change in *TFR* does not disentangle the change in fertility stock or quantum from the change in the timing of births or the change in the age distribution of fertility. In this paper, we have constructed the surface measure of fertility *(SMF)* which serves a more robust composite measure of fertility than *TFR* and allows decomposition of the change in *TFR* into the contribution of the change in fertility quantum and the change in age distribution of fertility or the timing of births. The change in 5MFprovides the true perspective of fertility transition as *SMF* is not influenced by the change in the age distribution of fertility. It is, therefore, suggested that *SMF* should be integrated into fertility monitoring frameworks along with *TFR* to provide a more nuanced picture of fertility transition by analysing the change in both fertility stock or quantum and age distribution of fertility or timing of births. The paper also reveals that the change in the age distribution of fertility in India has contributed to both slowing down the decrease in *TFR* relative to the decrease in *SMF* and hastening the decrease in *TFR* relative to the decrease in *SMF* which means that fertility transition depicted through the change in *TFR* is different from fertility transition depicted through the change in *SMF*. If the change in *SMF* is any indication, then fertility transition in the country has slowed down during the period 2011-2022. However, the change in *TFR* suggests that fertility transition in the country has accelerated during this period because of the change in the age distribution of fertility.

## Data Availability

All data produced in the present study are available upon reasonable request to the authors

